# Kangaroo mother care for preterm or low birth weight infants: A systematic review and meta-analysis

**DOI:** 10.1101/2022.09.14.22279053

**Authors:** Sindhu Sivanandan, Mari Jeeva Sankar

## Abstract

**Importance:** The Cochrane review (2016) on kangaroo mother care (KMC) demonstrated a significant reduction in the risk of mortality in low birth weight (LBW) infants. New evidence from large multi-center randomized trials has been available since its publication.

**Objective:** The objective of the systematic review was to compare the effects of KMC vs. conventional care and early (i.e., within 24 hours of birth) vs. late initiation of KMC on critical outcomes such as neonatal mortality.

**Methods:** Eight electronic databases, including PubMed, Embase, and Cochrane CENTRAL, until March 2022, were searched. All randomized trials comparing KMC versus conventional care or early vs. late initiation of KMC in LBW or preterm infants were included.

**Data extraction and synthesis:** The review followed the Preferred Reporting Items for Systematic Reviews and Meta-Analyses (PRISMA) guidelines and was registered with PROSPERO.

**Main outcomes and measures:** The primary outcome was mortality by 28 days of life. Other outcomes included severe infection, hypothermia, exclusive breastfeeding rates, and neurodevelopmental impairment. Results were pooled using fixed-effect and random-effects meta-analyses in RevMan 5.4 or Stata 15.1 (StataCorp, College Station, TX).

**Results:** In total, 31 trials with 15,559 infants were included in the review; 27 studies compared KMC with conventional care, while 4 compared early vs. late initiation of KMC. Compared to conventional care, KMC significantly reduced the risks of mortality (relative risk [RR] 0.68; 95% CI 0.53 to 0.86; 11 trials, 10505 infants; high certainty evidence) at discharge or 28 days of age and severe infection till the latest follow-up (RR 0.85, 95% CI 0.79 to 0.92; 9 trials; moderate certainty evidence). On subgroup analysis, KMC provided for a duration of at least 8 hours per day had more significant benefits compared to lesser duration KMC. Studies comparing early vs. late-initiated KMC demonstrated a significant reduction in neonatal mortality (RR 0.77, 95% CI 0.66 to 0.91; 3 trials, 3693 infants; high certainty evidence) and clinical sepsis till 28-days (RR 0.85, 95% CI 0.76 to 0.96; 2 trials; low certainty evidence) favoring early initiation of KMC.

**Conclusions and Relevance:** The review provides updated evidence on the effects of KMC on mortality and other critical outcomes in low birth weight infants. The findings suggest that KMC should preferably be initiated within 24 hours of birth and provided for at least 8 hours daily.

## INTRODUCTION

Prematurity (gestational age <37 weeks) and low birth weight (LBW, birth weight <2,500 g) are important causes of neonatal and infant mortality and long-term neurodevelopmental disability.^1^ Low- and middle-income countries (LMIC) have the highest burden of preterm and LBW infants. Kangaroo mother care (KMC) is a simple and cost-effective intervention that has been shown to reduce neonatal mortality and the risk of infection in LBW infants.^2^ The Cochrane review on KMC, published in 2016, included 21 studies involving 3042 infants and demonstrated a significant reduction in the risk of mortality and severe infection at the latest follow-up in LBW infants.^3^

New evidence from large multi-country and community-based randomized trials is available after the publication of the Cochrane review.^4,5^ Moreover, a few of them have examined the effect of early KMC, i.e., KMC initiated within the first 24 hours of delivery – often before stabilization of the infants.^5,6^ The timing of initiation of KMC is critical because KMC is usually commenced after the infant is stabilized. The World Health Organization (WHO) guidelines also recommend the initiation of KMC after clinical stabilization. However, stabilization of preterm/LBW neonates may take hours to days, depending upon the gestation, birth weight, and general condition at birth. Indeed, the median age at initiation of KMC in the facility-based studies included in the Cochrane review varied from 3 to 24 days. KMC initiated after 3 days of life would not naturally reduce the risk of deaths occurring in the first 3 days, which account for about 62% of total neonatal deaths^7^. The efficacy and safety of early initiation of KMC – within 24 hours or before stabilization – are unknown.

This systematic review aims to compare the effects of (a) KMC with conventional care and (b) early initiation, i.e., KMC within 24 hours of age, with late initiation of KMC on neonatal and infant mortality and severe morbidities among LBW infants. The updated review would provide critical evidence for policymakers and other stakeholders and help formulate clinical practice guidelines.

## METHODS

### Inclusion and exclusion criteria

The review included randomized and cluster-randomized trials that compared KMC with conventional care or early-initiated (i.e., in the first 24 hours after birth) KMC with late-initiated KMC among LBW or preterm infants, irrespective of the duration of KMC, infant stability at enrolment, study setting, and breastfeeding patterns. Trials reported as only abstracts were included if sufficient information on study methods was available to assess the eligibility and the risk of bias. We excluded quasi-randomized and crossover trials, studies evaluating KMC among term infants or those with birthweight >2500 g, and studies assessing KMC on only physiological parameters, pain scores, maternal mental health, infant colic, or during neonatal transport or as a part of a package of interventions.

### Search strategy

We systematically reviewed the relevant publications by searching the electronic databases of MEDLINE (1966-March 2022) via PubMed and OVID, Cochrane Central Register of Controlled Trials (CENTRAL, The Cochrane Library, Issue 1, March 2022), EMBASE (1988-March 2022), CINAHL (1981-March 2022), and the databases PsycINFO, AMED, EMCARE, BNI from inception till March 2022. We used the search terms “kangaroo care,” “kangaroo mother care,” “skin to skin care,” and “neonates or infants” in the search strategy. The search was initially conducted until March 2021 (for the presentation of review findings to the WHO Guideline Development Group of the upcoming guidelines on the care of LBW infants); the search was then updated till March 2022. The search strategy, search results, and the definitions used in the review are provided in **Supplement 1**. We also searched the databases of clinical trials and reference lists of retrieved articles for eligible studies.

### Outcomes

The primary outcome was mortality by day 28 of life. Other outcomes were mortality by 6-12 months of age, severe infections, infant growth, neurodevelopment, hypothermia, length of hospital stay, readmission to hospital, and exclusive breastfeeding at discharge and one and six months of age.

### Data extraction

The two review authors (SS and MJS) extracted data using a standardized and pre-tested data abstraction form. The data included study characteristics, sample size, details on KMC initiation, duration, breastfeeding, time of hospital discharge, study setting (hospital or community), outcomes including neonatal mortality, hypothermia, sepsis, re-hospitalization, rates of exclusive breastfeeding, and weight gain. Discrepancies, if any, were resolved by mutual discussion between the reviewers.

### Quality assessment and statistical analysis

The review authors independently evaluated the quality of studies using Cochrane’s Risk of Bias-1 tool, extracted data, and synthesized the effect estimates – relative risks (RR) or mean difference (MD) – using RevMan 5.4 or Stata 15.1 (StataCorp, College Station, TX). The RR and 95% confidence intervals (CI) were calculated based on the extracted frequencies and denominators. Results were pooled using fixed-effect meta-analyses using the Mantel-Haenszel method. The heterogeneity of the pooled studies was assessed using the test of homogeneity of study-specific effect sizes and the I^2^ statistic, in addition to visual confirmation from forest plots. If substantial heterogeneity was detected, the reasons for heterogeneity were explored. If there was no critical clinical or methodological heterogeneity among the studies, we pooled their results using the random-effects model. We evaluated the likelihood of potential publication bias using funnel plots.

The Grading of Recommendations Assessment, Development, and Evaluation (GRADE) approach^8^ was used to assess the quality of evidence for critical outcomes such as mortality at discharge, severe infection/sepsis at the latest follow-up, weight gain, exclusive breastfeeding, and neurodevelopmental outcomes. Evidence from randomized controlled trials was considered high quality; still, it could be downgraded by one or two levels for serious and very serious limitations, respectively, based on the risk of bias, imprecision, inconsistency, indirectness of study results, and publication bias. The review followed the Preferred Reporting Items for Systematic Reviews and Meta-Analyses (PRISMA) guidelines and was registered in PROSPERO (CRD42021240336).

### Planned subgroup analyses

For the comparison of KMC vs. conventional care, we performed subgroup analyses according to different gestational and birth weight categories and by median duration KMC in hours (<2 hours, 2-8 hours, 8-16 hours, and >16 hours); time of initiation of KMC - early (≤24 hours of life) vs. late initiation; stable vs. unstable neonates; setting - facility vs. community settings; and countries (high income vs. LMIC settings).

## RESULTS

Of the 3458 records identified from the database and bibliographic searches, 31^4-6,9-36^ studies enrolling 15,559 infants were included in the review (**Figure 1**); 25 studies were conducted in low-or middle-income countries (two from multiple countries^5,14^) while 7 were conducted in high-income countries^12,20,24,27,30,31,35^ (Appendix). Twenty-seven studies compared KMC with conventional care, while four had compared early with late initiation of KMC.^5,6,24,26^ KMC was initiated in the health facility in 29 studies and at home (community) in 2 trials.^4,11^ While the sample sizes of earlier hospital-based studies ranged from 28 to 777, the most recent facility-based study – WHO iKMC study^5^ – had a sample size of 3211. Of the two community-based studies, one trial had enrolled around 8400 infants.^4^ Only six studies included infants with birthweight < 1500 g.^12,13,19,29,31,35^ The characteristics of included studies are provided in **Table 1**. Figure 2 depicts the risk of bias in the included studies in specific domains. Many studies had an unclear or high risk of selection bias (due to a lack of information on allocation concealment) and detection bias (because the outcomes assessors were not masked to the intervention group).

**Table 1:** KMC versus conventional newborn care - characteristics of included studies.

| **S. No** | **Author, Year** | **Country** | **Stabilization status** | **Intervention group** | **Control group** | **Age at initiation of KMC (days)** | **KMC duration (hours per day)** | **Last follow-up** | **Schema for follow-up** |
| --- | --- | --- | --- | --- | --- | --- | --- | --- | --- |
| 1. | Ali 2009 | India | Stable | KMC in facility | Conventional warmer or cots | 4.7 | 6.3 ± 1.5 (range 4 to 12) | 6 months corrected age | Weekly until 40 weeks' postmenstrual age, fortnightly until 3 months corrected age, and monthly until 6 months corrected age |
| 2. | Alisjahbana 1998 | Indonesia | Stable | KMC in community | Conventional home care | - | - | 4 weeks after discharge | Weekly for 4 weeks after discharge |
| 3. | Archarya 2004 | Nepal | Stable | KMC in facility | Conventional warmer or cots | - | 6 | In-hospital | Not reported |
| 4. | Blaymore Bier 1996 | USA | Stable | KMC in facility | Conventional clothed | 29 | - | 6 months after hospital discharge | At 1, 3, and 6 months after hospital discharge |
| 5. | Boo 2007 | Malaysia | Stable | KMC in facility | Conventional NICU | 25 | 1 | In-hospital | Not reported |
| 6. | Cattaneo 1998 | Ethiopia | Stable | KMC in facility | Conventional open cribs, incubator or warmer | 10 | 20 | 30 days postnatal age | 4 times: at 3, 10, 20, and 30 days, and as usually scheduled at each hospital afterward |
| 7. | Charpak 1997 | Columbia | Stable | KMC in facility | Conventional incubator | 4 | 24 | 1 year and 20 years for a subset of enrolled subjected | At least once a week until 40 weeks' postmenstrual age; then monthly up to 3 months' corrected age, every 6 weeks until at least 6 months' corrected age, and every third month until 12 months' corrected age |
| 8. | Damala 2016 | India | Stable | KMC in facility | Conventional warmer or cots | - | 13-14 | Until 2.5 kg weight | After discharge, babies were followed up twice a week for the first week and weekly till babies reached 2.5 kg. |
| 9. | Eka Pratiwi 2009 | Indonesia | Stable | KMC in facility | Conventional warmer or cots | 1 | 10.0 ± 1.8 | In-hospital | - |

**Figure 1:**
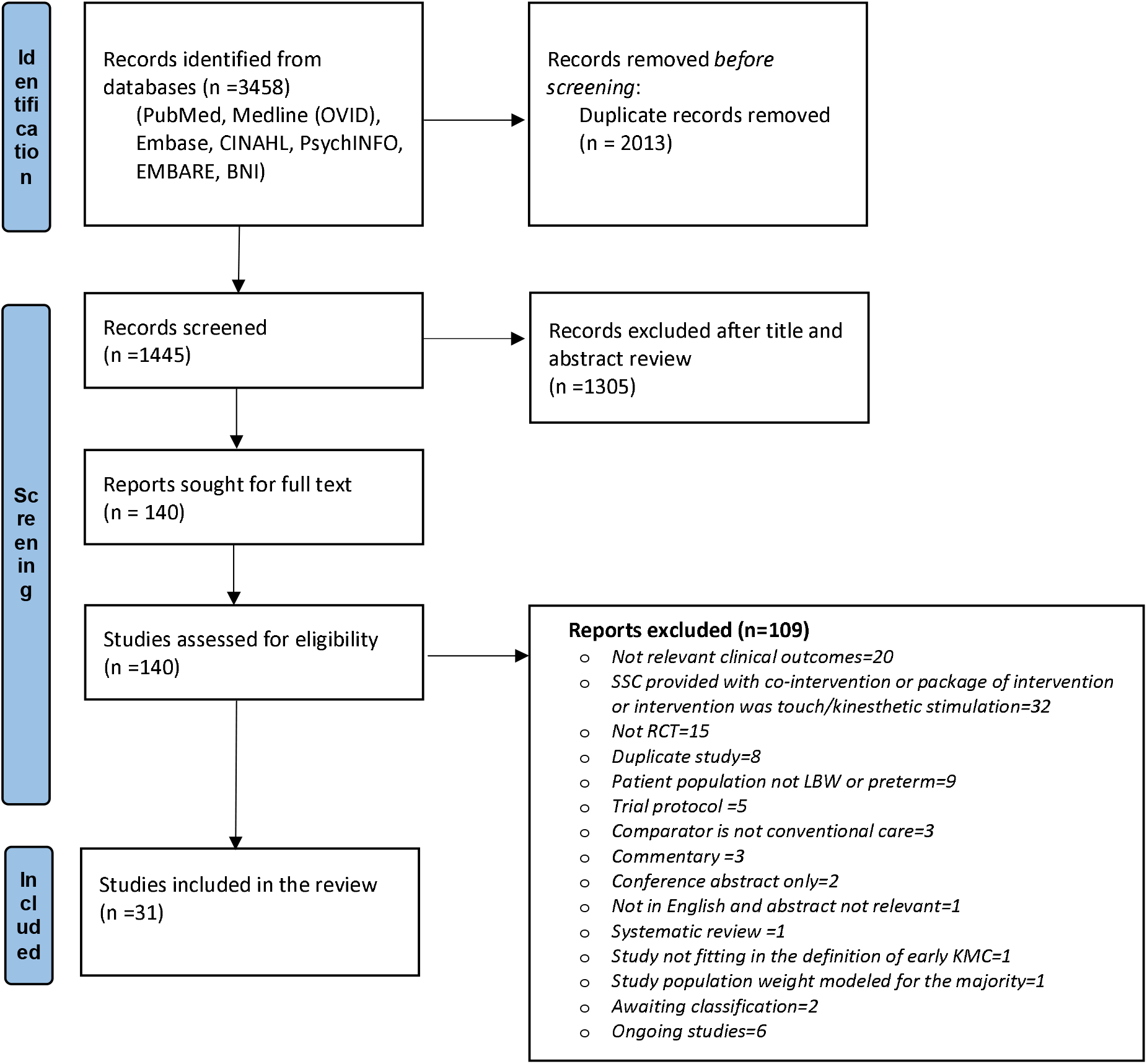
Flow chart of search results (adapted from PRISMA 2009 flow diagram)

**Figure 2:**
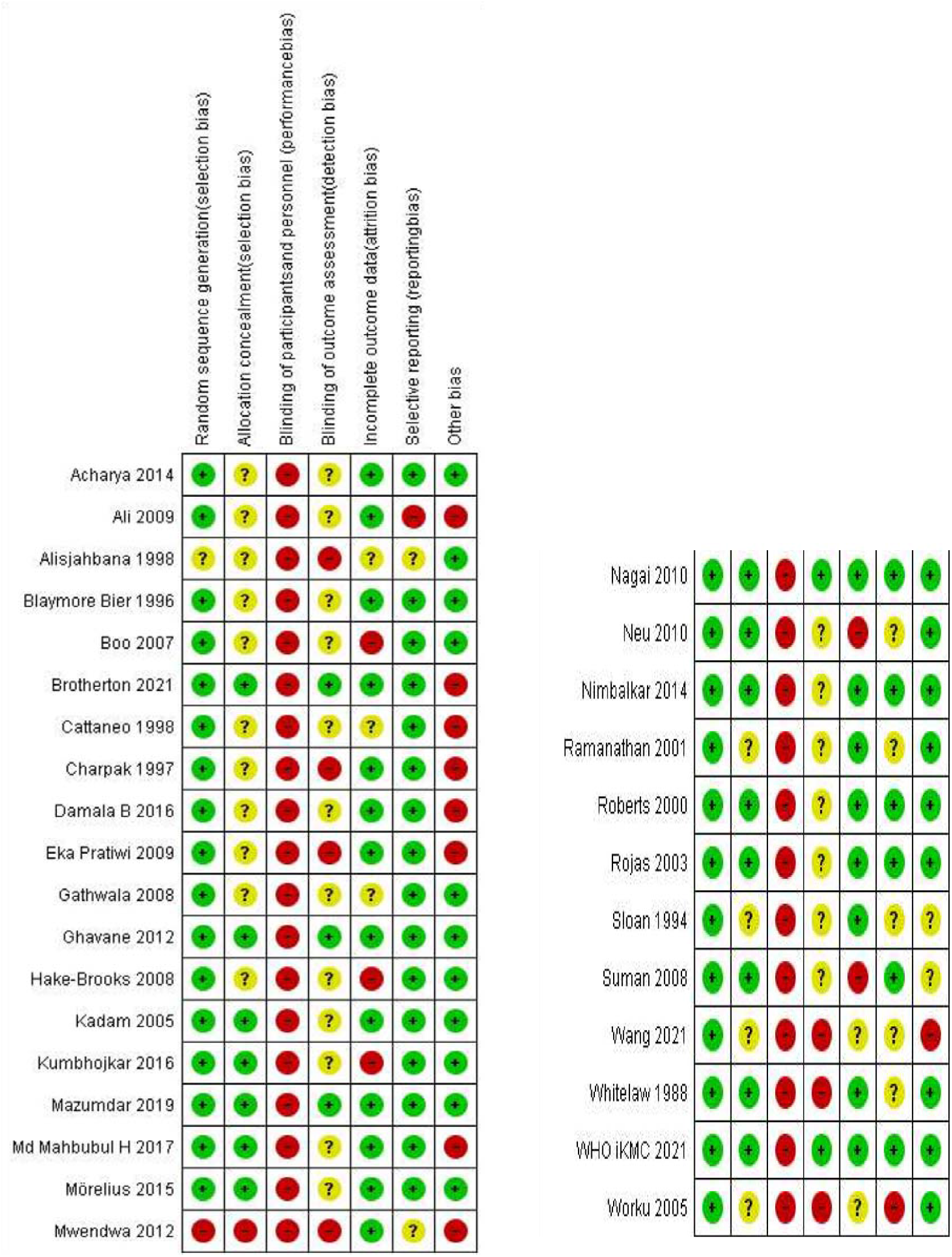
Risk of bias in included studies.

### KMC versus conventional newborn care

The comparison included 27 studies that enrolled 11,956 infants (Table 1). All but one study enrolled infants after stabilization (variably defined in different studies as cardiorespiratory stability, off oxygen or any form of respiratory support, or off intravenous fluids). KMC was started within 24 hours after birth in 2 studies, between 1 and 7 days in 10 studies, and after 7 days in 12 studies (3 studies did not report the time of initiation). The duration of KMC was <8 hours in 9 studies, 8-16 hours in 9 studies, and >16 hours in 4 studies (5 studies did not report the duration).

Pooled analysis revealed a 32% reduction in mortality by discharge or 40 weeks of postmenstrual age (PMA) or 28 days after birth (risk ratio [RR] 0.68; 95% confidence interval [CI] 0.53 to 0.86; I^2^ =0%; 12 studies; 10505 infants; high certainty evidence; Figure 3). The funnel plot did not show any evidence of a potential publication bias (eFigure 1 in **Supplement 2**). On subgroup analysis, mortality by discharge or 28 days was reduced for infants with gestational age ≤34 weeks as well as >34 weeks, weights ≤2000 g or >2000 g at birth/enrolment, for a daily duration of KMC of at least >8 hours per day (eFigure 2 in **Supplement 2)**, both in facility and community settings and with KMC initiated within 24 h after birth or later. Four studies had reported mortality by 6 months of age. There was 25% reduction in mortality (RR 0.75; 95% CI 0.62 to 0.92; high certainty of evidence).

**Figure 3:**
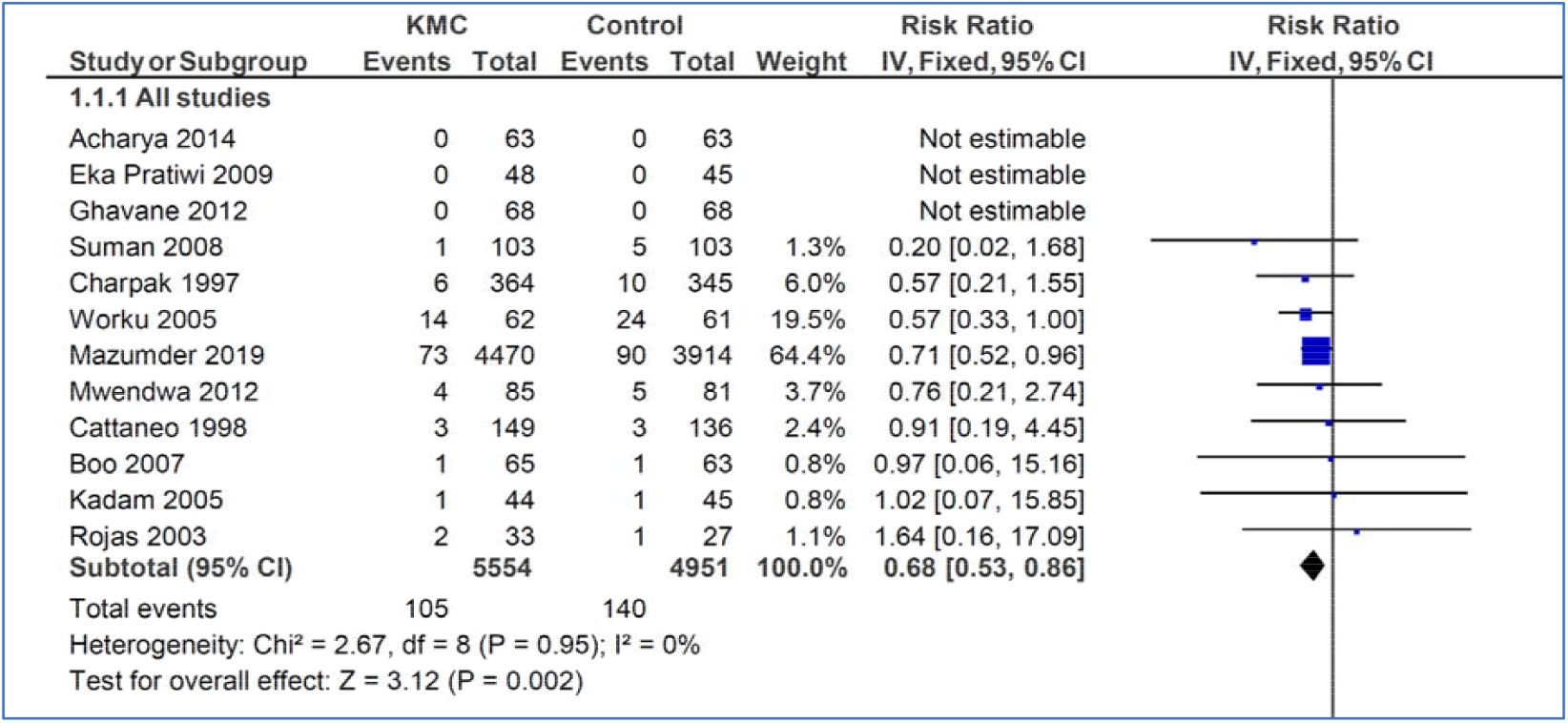
KMC vs. conventional care - forest plot of mortality by discharge or 28 days of life.

There was a 15% reduction in severe infection/sepsis at the latest follow-up (RR 0.85, 95% CI 0.79 to 0.92; 9 trials, 9847 infants; moderate certainty evidence). KMC also reduced the risk of hypothermia (RR 0.32, 95% CI 0.19 to 0.53; 11 trials, 1169 infants; moderate-certainty evidence). Infants in the KMC arm had a higher gain in anthropometric parameters, namely weight gain per day and length and head circumference gain per week (Table 2). The exclusive breastfeeding rates were higher at discharge/28 days of life (RR 1.48, 95% CI 1.44 to 1.52; 9 trials, 9983 infants, very low certainty evidence), but there was no difference at 1-3 months of age. There were no significant differences between KMC infants and controls in the Griffith Quotients or the risk of cerebral palsy at 12 months of corrected age^37^ or IQ scores at 20 years of age.

**Table 2:** KMC vs. conventional newborn care: key outcomes.

| Outcome and subgroup | Studies | N | Effect estimate |
| --- | --- | --- | --- |
| Mortality by discharge or 40 weeks' PMA or 28 days of age | 12 | 10,505 | 0.68 [0.53 to 0.87] |
| • Health facility setting | 11 | 2121 | 0.62 [0.41 to 0.94] |
| • Community settings | 1 | 8384 | 0.71 [0.52 to 0.96] |
| Mortality 6 months follow-up | 4 | 8031 | 0.75 [0.62 to 0.92] |
| • Health facility setting | 3 | 1047 | 0.74 [0.44 to 1.23] |
| • Community settings | 1 | 6984 | 0.76 [0.61 to 0.95] |
| *Severe infection\*/sepsis at latest follow-up* | 9 | 9847 | 0.85 [0.79, 0.92] |
| • Health facility setting | 8 | 1463 | 0.50 [0.36, 0.69] |
| • Community settings* | 1 | 8384 | 0.89 [0.82, 0.97] |
| *Hypothermia by discharge or by 40-41 weeks' PMA or 28 days follow-up* | 11 | 1169 | 0.32 [0.19, 0.53] |
| Exclusive breastfeeding at discharge or at 28 days of age | 9 | 9983 | 1.48 [1.44, 1.52] |
| • Health facility setting | 8 | 1599 | 1.18 [1.10, 1.27] |
| • Community settings | 1 | 8384 | 1.54 [1.49, 1.59] |
| Exclusive breastfeeding at 1 to 3 months' follow-up | 7 | 8139 | 1.39 [0.99, 1.97] |
| Weight gain at latest follow-up (g/d) | 11 | 1198 | 4.08 [2.30, 5.86] |
| Length gain at latest follow-up (cm/week) | 3 | 377 | 0.21 [0.03, 0.38] |
| Head circumference gain at latest follow-up (cm/week) | 5 | 652 | 0.18 [0.09, 0.27] |
| Cerebral palsy at 12 months' corrected age | 1 | 588 | 0.65 [0.21, 2.02] |
| Severe disability at 20 years | 1 | 264 | 0.34 [0.09, 1.24] |
| Neurodevelopmental outcomes at 12 months of age using BSID-III |  |  |  |
| Cognitive score | 1 | 516 | 0.21 [-1.84, 2.26] |
| Language scores | 1 | 516 | -0.91 [-2.46, 0.64] |
| Motor scores | 1 | 516 | -0.85 [-2.65, 0.95] |

*\*In community settings, the diagnosis of sepsis or severe infection was based on the World Health Organization (WHO) definition of possible serious bacterial infection. PMA; postmenstrual age, BSID-III; Bayley Scales of Infant Development-III*

### Early-initiated versus late-initiated KMC

The evidence was derived from 4 studies that enrolled 3603 infants. One study was done in a high-come country (Sweden), two were done in low-income countries (Madagascar and The Gambia), and one was a multi-country study conducted in LMICs (Ghana, India, Malawi, Nigeria, and Tanzania). All studies were conducted in health facilities. Infant stability at enrolment, duration of KMC achieved, and time of initiation of KMC in the included studies are provided in **Table 3**. In two studies (Mörelius et al.^24^ and WHO iKMC^5^), KMC was initiated in the delivery room. Brotherton et al.^6^ enrolled moderately unstable infants in the early KMC arm and stable infants after >24 h of admission in the control arm. Nagai et al. began KMC within 24 hours of birth in the early arm and after 24 hours in the late arm.

**Table 3:** Early- vs. late-initiated KMC – characteristics of included studies.

| **S. No** | **Author, Year** | **Inclusion criteria** | **Exclusion criteria** | **Intervention:**<br>**Early KMC as planned/**<br>**as achieved** | **Control:**<br>**Late KMC as planned/as**<br>**achieved** |
| --- | --- | --- | --- | --- | --- |
| 1 | WHO iKMC 2021 | All infants 1.0 and 1.799 kg, regardless of gestational age, type of delivery, or singleton or twin status (irrespective of clinical stability). | Infants who were unable to breathe spontaneously by 1 hour or who had a major congenital malformation | Immediately after birth;<br>**Median initiation time of 1.3 hrs after birth** | KMC began after the neonate recovered from preterm birth complications and was at least 24 hours old;<br>Median initiation time 53.6 h after birth |
| 2 | Brotherton 2021 | Birth weight <2000g and age 1-24 hours | Stable and severely unstable neonates excluded. Triplets, major congenital malformations; severe jaundice; seizures, lack of study bed | KMC initiated <24 h after admission;<br>**Median initiation time 13.6 h** | KMC once stable at >24 h after admission;<br>Median initiation time 104.5 h |
| 3 | Mörelus 2015 | Vaginally born singleton preterm infant (GA 32–35 weeks) (irrespective of clinical stability) | Infants with congenital malformations and severely unstable infants | Continuous skin-to-skin contact, beginning in the delivery room;<br>**Median initiation time not provided** | KMC began in the NICU;<br>On day two, both groups were practicing KMC |
| 4 | Nagai 2010 | birth weight under 2500 g, age less than 24 h post-birth, no serious malformation and relatively stable clinical condition | Apnea and intravenous infusion | KMC begun soon as possible, within 24 h post-birth;<br>**Median initiation time 19 hours (IQR 13.00–23.00)** | KMC begun after complete stabilization (generally after 24 h post-birth)<br>Median initiation time 28.5 hours (IQR 25–40) |

Early-initiated KMC showed a significant reduction in the risks of mortality by 28 days of age (RR 0.78, 95% CI 0.66 to 0.92; 3 trials, 3533 infants, high certainty evidence; **eFigure 3)**, clinical sepsis till 28-day follow-up (RR 0.85, 95% CI 0.76 to 0.96; **Table 4;** low certainty evidence) and hypothermia by discharge or at 28 days (RR 0.74, 95% CI 0.61 to 0.90; high certainty evidence), and improvement in exclusive breastfeeding at discharge (RR 1.1.2, 95% CI 1.10 to 1.19; moderate certainty evidence). There was also a decrease in the length of hospital stay (**Table 4**).

**Table 4:** Early- vs. late-initiated KMC – critical outcomes.

| **Outcome** | **Studies** | **Number of participants** | **Pooled relative risk**<br>**(95% confidence interval)** |
| --- | --- | --- | --- |
| Mortality by 28 days of life | 3 | 3533 | 0.78 (0.66 to 0.92) |
| Mortality at 6 months of age | 1 | 72 | 1.0 (0.15 to 6.72) |
| Sepsis till 28 days | 2 | 3415 | 0.85 (0.76 to 0.96) |
| Exclusive breastfeeding at discharge | 3 | 3464 | 1.12 (1.07 to 1.16) |
| Exclusive breastfeeding at 28 days of age | 3 | 2841 | 1.01 (0.98 to 1.04) |
| Hypothermia at discharge or by 28 days | 3 | 3553 | 0.74 (0.61 to 0.90) |
| Weight gain at 28-day follow-up (g/d) | 1 | 204 | MD -2.20 (-5.26 to 0.86) |
| Nosocomial sepsis |  |  |  |
| Clinical sepsis | 2 | 3415 | 0.85 (0.75 to 0.95) |
| Culture-positive sepsis | 1 | 279 | 1.53 (0.44 to 5.31) |
| Re-admission to hospital at 4 weeks of age | 1 | 73 | 1.95 (0.18 to 20.5) |
| Length of hospital stay (days) | 3 | 3498 | -0.30 (-0.31 to -0.29) |

*MD, mean difference*

On subgroup analysis, there was evidence of a reduction in 28-day mortality for infants with GA ≤34 weeks and BW ≤2000, but there was little data for infants >34 weeks and weighing >2000 g at birth. The mortality reduced with a duration of KMC of at least >16 hours per day, with little data for daily KMC duration of <8 hours or 8-16 hours per day.

### Quality of the evidence

For the comparison of KMC vs. conventional newborn care, the certainty of the evidence was assessed as high for neonatal mortality and moderate for sepsis/severe infection and hypothermia (eTable 1 in **Supplement 2)**. For early vs. late-initiated KMC, the certainty of the evidence was high for neonatal mortality and hypothermia, moderate for exclusive breastfeeding at discharge, and low for nosocomial clinical sepsis (eTable 2 in **Supplement 2)**. A few outcomes, such as weight gain, breastfeeding, and length of hospital stay, showed a high degree of heterogeneity, partly due to clinical and methodological heterogeneity among the studies (varied definitions of hypothermia and time-points of assessment; different methods of breastfeeding assessment, etc.).

## DISCUSSION

The systematic review results showed a significant reduction in mortality at discharge or 28 days of age, severe infection or sepsis, and hypothermia at the latest follow-up in LBW infants receiving KMC in the health facility or at home. KMC increased the weight and length gain but did not improve the exclusive breastfeeding rates at the latest follow-up; neurodevelopmental outcomes at 12 months were also not different between the KMC and conventional care groups. Compared to delayed initiation (> 24 hours) of KMC, early-initiated KMC (< 24 hours) results in a 33% reduction in mortality by 28 days, 25% and 36% reduction in the risks of clinical sepsis and hypothermia, respectively, and a 12% improvement in exclusive breastfeeding at discharge.

Three recent systematic reviews have examined the effect of KMC compared to conventional care on infant clinical outcomes.^3,38,39^ The Cochrane review in 2016 found 21 studies enrolling 3042 LBW infants.^3^ Our updated search used a similar search strategy and inclusion criteria to the Cochrane review. We found another 10 studies that provided data on 12,517 additional infants with similar gestation and birth weight range. The Cochrane review reported a similar decrease in mortality at discharge or 40 weeks of postmenstrual age (RR 0.60, 95% CI 0.39 to 0.92; 8 trials, 1736 infants) and similar effects on infection, hypothermia, and anthropometry. However, the certainty of the evidence was graded as moderate to very low in the Cochrane review. The addition of information from 12,000-odd infants has improved the precision and certainty of the evidence of the critical outcomes in the current study. In 2020, a systematic review of 416 preterm neonates reported that KMC significantly reduced apneic events in preterm neonates ^39^ Another review^38^in 2019 reported that KMC had a significant positive impact on growth and breastfeeding rates in very low birth weight (VLBW) neonates.

We investigated the effect of mean duration KMC in hours and prespecified three categories (<8 hours, 8-16 hours, and >16 hours). The effects on mortality were comparable in the >16 h and 8-16 h groups, but there was insufficient data in the <8 hours group. The Cochrane review (2016) explored the effects of the duration of KMC in three different categories; <2 hours and 6-15 hours, and >20 hours per day, and found benefits only when KMC was done for 20 hours or more. We found beneficial effects of KMC in prespecified subgroups of ≤2.0 kg and >2.0 kg, infants with gestational age ≤34 and >34 weeks at birth. The two community-based studies that enrolled infants at home also showed significant benefits on mortality. We found no additional trials (other than that included in the Cochrane review) that compared KMC with conventional care in unstable infants.

Only one systematic review – the Cochrane review published in 2016 – has evaluated the effects of early vs. late initiation of KMC in LBW infants. It also used a cut-off of 24 hours to define early initiation but found only one study of 73 relatively stable LBW infants.^26^ Our review included three additional studies that recruited 3530 preterm/LBW infants and found significant beneficial effects with early initiation of KMC. ^5,6,24^

The results of our review have substantial implications for policymaking, particularly in low-and middle-income countries (LMIC). First, KMC should be provided to all low birth weight and preterm infants irrespective of the settings – both health facilities and at home. Second, given the probable dose-effect response, KMC should preferably be practiced for at least 8 hours daily for optimal benefits. Third, KMC should be initiated within the first 24 hours of life – irrespective of whether the infant is stabilized. Indeed, our findings have helped make recommendations on KMC in the upcoming WHO guidelines on the care of preterm neonates.

The strengths of the current review include a comprehensive and systematic search of the literature with the updated evidence till March 2022. Compared to the existing Cochrane reviews on KMC, this review identified additional studies that had enrolled almost 13000 LBW infants, which resulted in high precision of estimates and improved the certainty of the evidence. The review had some limitations too. The included studies were not blinded, though outcome assessors were blinded in many studies. However, the risk of bias in the included studies was generally low, and the certainty of the evidence for the primary outcomes was moderate to high. Some subgroup analyses had only a few studies or a small number of enrolled infants that precluded making firm conclusions.

Our findings support the practice of KMC for all preterm and LBW infants as soon as possible after birth and for at least eight hours per day regardless of the infant’s gestational age, birth weight, or stability. Future research should focus on overcoming barriers and facilitators to large-scale implementation of KMC in both facility and community settings. Data on long-term neurodevelopmental outcomes are also needed.

## Supporting information

Search strategy

Summary of Findings Table and additional data

## Data Availability

All data produced in the present work are contained in the manuscript

## ACKNOWLEDGMENTS

We acknowledge the support and guidance provided by Dr. Rajiv Bahl, Dr. Karen Edmond, and Dr. Shuchita Gupta from the WHO, Geneva, in finalizing the protocol and interpreting the results.

## Key Points

### Question

What are the effects of kangaroo mother care (KMC) and early initiation of KMC (within 24 hours of life) on critical outcomes like neonatal mortality in low birth weight infants?

### Findings

In this meta-analysis of 31 randomized trials (15559 infants), KMC significantly reduced the risk of neonatal mortality (relative risk [RR] 0.68; 95% CI 0.53 to 0.86) when compared to conventional care. Similarly, compared to late initiation of KMC, early-initiated KMC reduced the risk of neonatal mortality by 33% (RR 0.77; 95% CI 0.66 to 0.91).

### Meaning

The findings provide updated evidence on the effects of KMC on mortality and other outcomes in LBW infants and suggest that KMC should preferably be initiated within 24 hours of birth.

