## Supplementary material for "Kangaroo mother care for preterm or low birth weight infants: A systematic review and meta-analysis": Search strategy

Supplement 1

**Definitions used in the review**

#### Panel 1: Definitions used in the review

**Kangaroo mother care:** Skin–to–skin contact between mother and newborn will be taken as the minimal core component of KMC. Additional components include frequent or exclusive breastfeeding and early discharge

**'Early initiated' KMC:** KMC provided immediately or in the first 24 hours of birth

**'Late’ initiation of KMC:** KMC provided after 24 hours of birth

**KMC in stable babies:** KMC provided to babies who are considered ‘stable’ before enrolment as defined by the study authors

**KMC in unstable babies:** KMC provided to babies who were not considered stable before enrolment by the study authors

**Outcomes and their definitions**

| **Primary outcome** | **Definition** | **Timepoint of ascertainment** |
| --- | --- | --- |
| Mortality | All-cause mortality | At discharge, at 28 d, and latest follow up |
| Hypothermia | As defined in individual studies | In-hospital or at home during study period |
| Severe infection/sepsis | As defined in individual studies or determined by the culture of bacteria or fungus from blood, cerebrospinal fluid, urine, or from a normally sterile body space OR Any clinically defined infection, including possible serious bacterial infection (PSBI), diarrhea, pneumonia | At discharge, at 28 d, and latest follow up |
| Severe illness | As defined in individual studies | At discharge, at 28 d, and latest follow up |
| Infant growth | Weight, Length, Head circumference  (as mean, z score < 2 SD or < 10^th^ centile for the index population) | At discharge, at 28 d, and latest follow up |
| Neurodevelopment | Neurodevelopment or educational scores measured using validated assessment tools. | At 12m, and latest follow up |
| Neurodevelopmental disability | Severe neurodevelopmental disability, defined as anyone or a combination of the following: non-ambulant cerebral palsy, auditory and visual impairment, neurodevelopmental impairment | At 12m, and latest follow up |

Search terms and search results

**PubMed- 31^st^ March 2021**

| **Search** | **Query** | **Results** |
| --- | --- | --- |
| #8 | Search: **#1 AND #2 AND #7** Sort by: **Most Recent** | 459 |
| #7 | Search: **#4 AND #5** Sort by: **Most Recent** | 2,636,074 |
| #6 | Search: **#1 AND #2 AND #3** Sort by: **Most Recent** | 478 |
| #5 | Search: (epidemiology OR case control study OR cohort analysis OR cross-sectional study OR case study OR follow up OR longitudinal study OR retrospective study OR prospective study OR observational study OR case control OR epidemiologic OR case referent OR case stud* OR case series OR cohort OR cross sectional OR follow up OR longitudinal OR retrospective* OR prospective* OR observational OR adverse effect OR interrupted time series OR correlational OR ecological stud* OR descriptive stud*). Sort by: Most Recent | 9,268,175 |
| #4 | Search: communit* OR home-base* OR homebase* OR county OR domiciliary OR developing OR disadvantaged OR facility OR home OR impoverished OR peripheral OR poor OR rural OR slum OR underdeveloped OR underserved unit* OR village* OR residence characteristics OR rural population OR developing countries Sort by: Most Recent | 7,419,770 |
| #3 | Search: (randomized controlled trial [pt] OR controlled clinical trial [pt] OR randomized [tiab] OR placebo [tiab] OR drug therapy [sh] OR randomly [tiab] OR trial [tiab] OR groups [tiab]) Sort by: Most Recent | 5,024,760 |
| #2 | Search: (((((((((infant[MeSH Terms]) OR (infant, low birth weight[MeSH Terms])) OR (infant, mortality[MeSH Terms])) OR (breastfeeding[MeSH Terms])) OR (Mother-Child Relations[MeSH Terms])) OR (infant care[MeSH Terms])) OR (length of stay[MeSH Terms])) OR (physical stimulation[MeSH Terms])) OR (weight gain[MeSH Terms])) OR ((infant, newborn[MeSH] OR newborn OR neonate OR neonatal OR premature OR low birth weight OR VLBW OR LBW or infan* or neonat*)) Sort by: Most Recent | 2,052,623 |
| #1 | Search: ((((((kangaroo care)) OR ("kangaroo care"[Text Word])) OR ("kangaroo mother care"[Text Word])) OR ("kangaroo mother method"[Text Word])) OR (skin-skin care[Text Word])) OR (skin-to-skin contact) Sort by: Most Recent | 1,913 |

**Embase 31^st^ March 2021**

| 1. | (kangaroo mother care OR kangaroo mother method OR kangaroo care OR skin-to-skin contact OR skin-to-skin care).ti,ab,af,if | 2,601 |
| --- | --- | --- |
| 2. | (infant, newborn OR newborn OR neonate OR neonatal OR premature OR very low birth weight OR low birth weight OR VLBW OR LBW OR Newborn OR infan* OR neonat*).ti,ab | 967,431 |
| 3. | (randomized controlled trial controlled clinical trial OR randomized OR placebo OR clinical trial OR randomly OR trial OR clinical trial).ti,ab,cf,cg,pt | 1,815,889 |
| 4. | *(#1 AND #2 AND #3)* | 428 |
| 5. | (communit* OR home base* OR homebase* OR county OR domiciliary OR developing OR disadvantaged OR facility OR home OR impoverished OR peripheral OR poor OR rural OR slum OR underdeveloped OR underserved unit* OR village* OR residence characteristics OR rural population OR developing countries).ti,ab,if | 3,530,921 |
| 6. | EPIDEMIOLOGY/ OR exp CASE CONTROL STUDY/ OR cohort analysis | 1,042,933 |
| 7. | CROSS-SECTIONAL STUDY/ OR CASE STUDY/ OR follow up OR LONGITUDINAL STUDY/ | 2,608,527 |
| 8. | RETROSPECTIVE STUDY/ OR PROSPECTIVE STUDY/ OR OBSERVATIONAL STUDY/ | 1,767,844 |
| 9. | (case control).ti,ab,cf,cg,pt | 171,831 |
| 10. | (epidemiologic).ti,ab,pt | 85,047 |
| 11. | (case referent).ti,ab,pt | 723 |
| 12. | (case stud*).ti,ab,pt | 132,190 |
| 13. | (case series OR cohort OR cross sectional OR follow up OR longitudinal OR retrospective* OR prospective* OR observational OR adverse effect OR (controlled before AND after) OR interrupted time series OR correlational OR ecological stud* OR descriptive stud*).ti,ab,pt | 4,641,879 |
| 14. | *5-13 (combined with OR)* | 5,932,137 |
| 15. | *(1 AND 2 AND 4 AND 14)* | 239 |

**CINAHL and Medline**

| 1. | (kangaroo mother care OR kangaroo care method OR skin-to-skin care OR skin-to-skin contact).ti,ab,af,mj | 2,184 |
| --- | --- | --- |
| 2. | (infant, newborn OR newborn OR neonate OR neonatal OR premature OR low birth weight OR VLBW OR LBW OR Newborn OR infan* OR neonat*).ti,ab,af,mj | 538,234 |
| 3. | (randomized controlled trial OR controlled clinical trial OR randomized OR placebo OR clinical trials as topic OR randomly OR trial OR PT clinical trial).ti,ab,af,mj | 610,826 |
| 4. | *(1 AND 2 AND 3)* | 315 |
| 5. | EPIDEMIOLOGY/ OR exp CASE CONTROL STUDY/ OR cohort analysis | 125,097 |
| 6. | CROSS-SECTIONAL STUDY/ OR CASE STUDY/ OR follow up OR LONGITUDINAL STUDY/ | 266,536 |
| 7. | RETROSPECTIVE STUDY/ OR PROSPECTIVE STUDY/ OR OBSERVATIONAL STUDY/ | 0 |
| 8. | (epidemiologic).ti,ab,pt | 17,488 |
| 9. | (case referent).ti,ab,pt | 552 |
| 10. | (case stud*).ti,ab,pt | 682,098 |
| 11. | (case series OR cohort OR cross sectional OR follow up OR longitudinal OR retrospective* OR prospective* OR observational OR adverse effect OR (controlled before AND after) OR interrupted time series OR correlational OR ecological stud* OR descriptive stud*).ti,ab,pt | 1,041,862 |
| 12. | *5-11 (combined with OR)* | 1,585,565 |
| 13. | (communit* OR home base* OR homebase* OR county OR domiciliary OR developing OR disadvantaged OR facility OR home OR impoverished OR peripheral OR poor OR rural OR slum OR underdeveloped OR underserved unit* OR village* OR residence characteristics OR rural population OR developing countries).ti,ab,hw | 948,279 |
| 14. | *(1 AND 2 AND 12 AND 13)* | 144 |

Medline (OVID) 31^st^ March 2021

| 1. | (kangaroo mother care OR kangaroo care method OR skin-to-skin care OR skin-to-skin contact).ti,ab,mm,mh,if,mj | 1,796 |
| --- | --- | --- |
| 2. | (infant, newborn OR newborn OR neonate OR neonatal OR premature OR low birth weight OR VLBW OR LBW OR Newborn).ti,ab,mm,mh,if,mj | 895,376 |
| 3. | (randomized controlled trial OR controlled clinical trial OR randomized OR placebo OR clinical trials as topic OR randomly OR trial OR PT clinical trial).ti,ab,mm,mh,if,mj | 1,415,815 |
| 4. | *(1 AND 2 AND 3)* | 315 |
| 5. | (community OR home based OR homebased OR county OR domiciliary OR developing OR disadvantaged OR facility OR home OR impoverished OR peripheral OR poor OR rural OR slum OR underdeveloped OR underserved unit OR village OR residence characteristics OR rural population OR developing countries).ti,ab,mm,mh,if,mj | 2,728,162 |
| 6. | (epidemiology OR case control study OR cohort analysis OR cross-sectional study OR case study OR follow up OR longitudinal study OR retrospective study OR prospective study OR observational study OR case control OR epidemiologic OR case referent OR case study OR case series OR cohort OR cross sectional OR follow up OR longitudinal OR retrospective OR prospective OR observational OR adverse effect OR interrupted time series OR correlational OR ecological study OR descriptive study).ti,ab,mm,mh,if,mj | 3,618,190 |
| 7. | *(1 AND 2 AND 5 AND 6)* | 177 |

Other databases: 31^st^ March 2021

| 1 | PsycINFO | (kangaroo mother care OR kangaroo care method OR skin-to-skin care OR skin-to-skin contact).ti,ab | 327 |
| --- | --- | --- | --- |
| 2 | PsycINFO | (infant, newborn OR newborn OR neonate OR neonatal OR premature OR low birth weight OR VLBW OR LBW OR Newborn).ti,ab | 39,101 |
| 3 | PsycINFO | (randomized controlled trial OR controlled clinical trial OR randomized OR placebo OR clinical trials as topic OR randomly OR trial OR PT clinical trial).ti,ab | 229,406 |
| 4 | PsycINFO | (community OR home based OR homebased OR county OR domiciliary OR developing OR disadvantaged OR facility OR home OR impoverished OR peripheral OR poor OR rural OR slum OR underdeveloped OR underserved unit OR village OR residence characteristics OR rural population OR developing countries).ti,ab | 695,732 |
| 5 | PsycINFO | (epidemiology OR case control study OR cohort analysis OR cross-sectional study OR case study OR follow up OR longitudinal study OR retrospective study OR prospective study OR observational study OR case control OR epidemiologic OR case referent OR case study OR case series OR cohort OR cross sectional OR follow up OR longitudinal OR retrospective OR prospective OR observational OR adverse effect OR interrupted time series OR correlational OR ecological study OR descriptive study).ti,ab | 644,952 |
|  | **PsycINFO** | ***(1 AND 2 AND 3)*** | **37** |
|  | **PsycINFO** | ***(1 AND 2 AND 4 AND 5)*** | **14** |
|  | AMED | (kangaroo mother care OR kangaroo care method OR skin-to-skin care OR skin-to-skin contact).ti,ab | 5 |
| 1 | EMCARE | (kangaroo mother care OR kangaroo care method OR skin-to-skin care OR skin-to-skin contact).ti,ab | 1,153 |
| 2 | EMCARE | (infant, newborn OR newborn OR neonate OR neonatal OR premature OR low birth weight OR VLBW OR LBW OR Newborn).ti,ab | 118,415 |
| 3 | EMCARE | (randomized controlled trial OR controlled clinical trial OR randomized OR placebo OR clinical trials as topic OR randomly OR trial OR PT clinical trial).ti,ab | 493,916 |
| 4 | EMCARE | (community OR home based OR homebased OR county OR domiciliary OR developing OR disadvantaged OR facility OR home OR impoverished OR peripheral OR poor OR rural OR slum OR underdeveloped OR underserved unit OR village OR residence characteristics OR rural population OR developing countries).ti,ab | 868,173 |
| 5 | EMCARE | (epidemiology OR case control study OR cohort analysis OR cross-sectional study OR case study OR follow up OR longitudinal study OR retrospective study OR prospective study OR observational study OR case control OR epidemiologic OR case referent OR case study OR case series OR cohort OR cross sectional OR follow up OR longitudinal OR retrospective OR prospective OR observational OR adverse effect OR interrupted time series OR correlational OR ecological study OR descriptive study).ti,ab | 1,240,198 |
|  | **EMCARE** | ***(1 AND 2 AND 3)*** | **148** |
|  | **EMCARE** | ***(1 AND 2 AND 4 AND 5)*** | **66** |
| 1 | BNI | (kangaroo mother care OR kangaroo care method OR skin-to-skin care OR skin-to-skin contact).ti,ab | 539 |
| 2 | BNI | (infant, newborn OR newborn OR neonate OR neonatal OR premature OR low birth weight OR VLBW OR LBW OR Newborn).ti,ab | 17,783 |
| 3 | BNI | (randomized controlled trial OR controlled clinical trial OR randomized OR placebo OR clinical trials as topic OR randomly OR trial OR PT clinical trial).ti,ab | 38,641 |
| 4 | BNI | (community OR home based OR homebased OR county OR domiciliary OR developing OR disadvantaged OR facility OR home OR impoverished OR peripheral OR poor OR rural OR slum OR underdeveloped OR underserved unit OR village OR residence characteristics OR rural population OR developing countries).ti,ab | 140,284 |
| 5 | BNI | (epidemiology OR case control study OR cohort analysis OR cross-sectional study OR case study OR follow up OR longitudinal study OR retrospective study OR prospective study OR observational study OR case control OR epidemiologic OR case referent OR case study OR case series OR cohort OR cross sectional OR follow up OR longitudinal OR retrospective OR prospective OR observational OR adverse effect OR interrupted time series OR correlational OR ecological study OR descriptive study).ti,ab | 105,504 |
|  | **BNI** | ***(1 AND 2 AND 3)*** | **54** |
|  | **BNI** | ***(1 AND 2 AND 4 AND 5)*** | **26** |

**Cochrane CENTRAL search terms**

Search terms "kangaroo care" OR "kangaroo mother care" OR "skin to skin care" OR "skin-skin care" OR kangaroo in Title Abstract Keyword - (Word variations have been searched)- 378 Trials matching

**UPDATED Search (31 March 2021 to 31 March 2022)**

**PubMed**

| #7 | Search: **#5 AND #6** | 59 |
| --- | --- | --- |
| #6 | Search: **#1 AND #2 AND #3 AND #4** | 545 |
| #5 | Search: ("2021/03/31"[Date - Create] : "2022/03/31"[Date - Create]) | 1,554,148 |
| #4 | Search: (epidemiology OR case control study OR cohort analysis OR cross-sectional study OR case study OR follow up OR longitudinal study OR retrospective study OR prospective study OR observational study OR case control OR epidemiologic OR case referent OR case stud* OR case series OR cohort OR cross sectional OR follow up OR longitudinal OR retrospective* OR prospective* OR observational OR adverse effect OR interrupted time series OR correlational OR ecological stud* OR descriptive stud*). | 10,118,843 |
| #3 | Search: communit* OR home-base* OR homebase* OR county OR domiciliary OR developing OR disadvantaged OR facility OR home OR impoverished OR peripheral OR poor OR rural OR slum OR underdeveloped OR underserved unit* OR village* OR residence characteristics OR rural population OR developing countries | 8,194,370 |
| #2 | Search: : (((((((((infant[MeSH Terms]) OR (infant, low birth weight[MeSH Terms])) OR (infant, mortality[MeSH Terms])) OR (breastfeeding[MeSH Terms])) OR (Mother-Child Relations[MeSH Terms])) OR (infant care[MeSH Terms])) OR (length of stay[MeSH Terms])) OR (physical stimulation[MeSH Terms])) OR (weight gain[MeSH Terms])) OR ((infant, newborn[MeSH] OR newborn OR neonate OR neonatal OR premature OR low birth weight OR VLBW OR LBW or infan* or neonat*)) | 2,159,091 |
| #1 | Search: ((((((kangaroo care)) OR ("kangaroo care"[Text Word])) OR ("kangaroo mother care"[Text Word])) OR ("kangaroo mother method"[Text Word])) OR (skin-skin care[Text Word])) OR (skin-to-skin contact) | 2,261 |

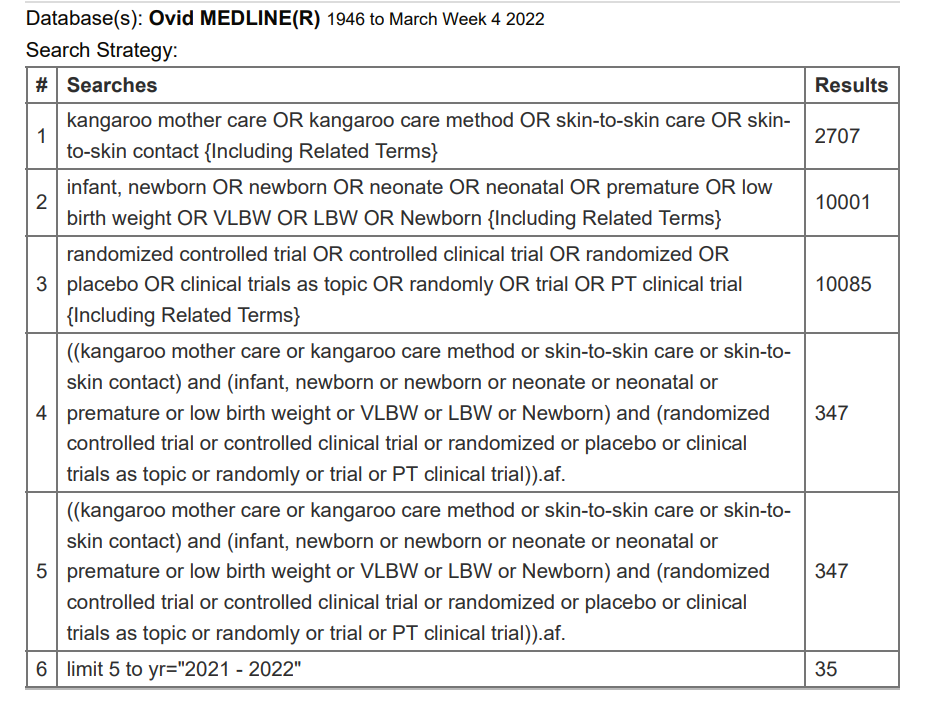

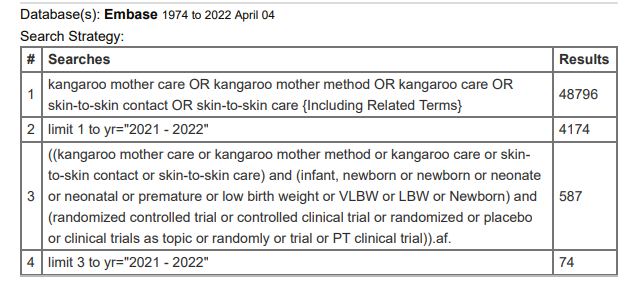

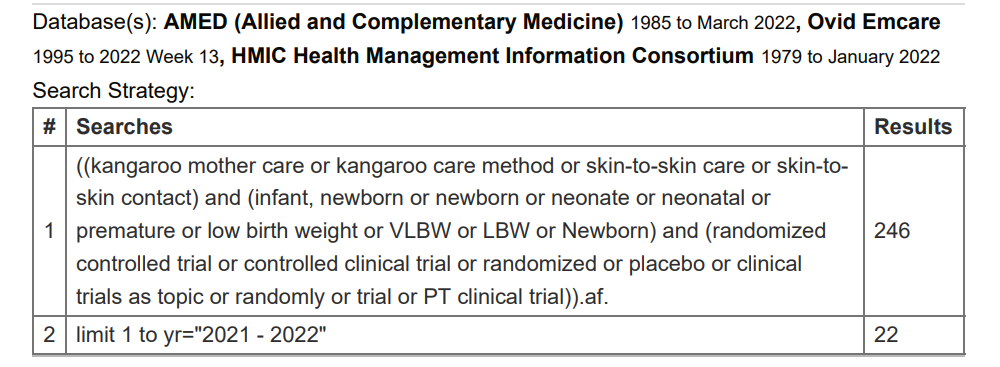
