## Supplementary material for "Kangaroo mother care for preterm or low birth weight infants: A systematic review and meta-analysis": Summary of Findings Table and additional data

**Supplement 2: eTables and eFigures**

**eTable 1: Summary of findings – KMC vs. conventional newborn care**

| **Summary of findings Table-1. Kangaroo mother care compared to conventional newborn care in preterm or low birthweight infants** | | | | | | | | |
| --- | --- | --- | --- | --- | --- | --- | --- | --- |
| ***Patient or population****: preterm or low birthweight infants*  ***Setting****: Hospital or community/home*  ***Intervention****: Kangaroo mother care*  ***Comparison****: Conventional newborn care* | | | | | | | | |
| ***Outcomes*** | ***№ of participants  (studies) Follow up*** | | | ***Certainty of the evidence (GRADE)*** | ***Relative effect (95% CI)*** | | ***Anticipated absolute effects*** | |
|  |  |  |  |  |  |  | ***Risk with conventional neonatal care*** | ***Risk difference with Kangaroo mother care*** |
| *Mortality by discharge or 40 weeks PMA or 28 days of age* | *10505 (12 RCTs)* | | | *⨁⨁⨁⨁ HIGH^a^* | ***RR 0.68*** *(0.53 to 0.87)* | | *28 per 1,000* | *9 fewer per 1,000*  *(from 13 fewer to 4 fewer)* |
| *Severe infection or sepsis till latest follow-up* | *9847 (9 RCTs)* | | | *⨁⨁⨁◯ MODERATE^b^* | ***RR 0.85*** *(0.79 to 0.92)* | | *215 per 1,000* | *32 fewer per 1,000 (45 fewer to 17 fewer)* |
| *Hypothermia by discharge or 40 weeks’ PMA or 28 days after birth* | *1169 (11 RCTs)* | | | *⨁⨁⨁◯ MODERATE ^c,d^* | ***RR 0.32*** *(0.19 to 0.53)* | | *257 per 1,000* | *175 fewer per 1,000 (from 208 fewer to 121 fewer)* |
| *Weight gain at latest follow-up (g/d)* | *1198 (11 RCTs)* | | | *⨁⨁◯◯ LOW^d,e^* | *-* | | *Mean weight gain at latest follow-up was 17 grams/day* | *MD 4.08 g/day higher (2.3 higher to 5.86 higher)* |
| *Exclusive breastfeeding* *at discharge or at 40 to 41 weeks' PMA or at 28 days of age* | *9983 (9 RCTs)* | | | *⨁◯◯◯ VERY LOW ^d,,f^* | ***RR 1.48*** *(1.44 to 1.52)* | | *546 per 1,000* | *262 more per 1,000 (from 240 more to 284 more)* |
| *Neurodevelopmental outcome at 12 months' using BSID-III (stable LBW infants)* | | *516*  *(1 RCT)* | *⨁⨁◯◯ LOW ^g,h,i^* | | | *Post-hoc equivalence testing using two one-sided tests of equivalence (TOST) demonstrated that composite scores for cognitive, language, and motor domains at 12 months among the study arms were statistically equivalent* | | |
| ******The risk in the intervention group*** *(and its 95% confidence interval) is based on the assumed risk in the comparison group and the* ***relative effect*** *of the intervention (and its 95% CI).*  ***CI:*** *Confidence interval;* ***RR:*** *Risk ratio;* ***MD:*** *Mean difference; PMA- Postmenstrual age* | | | | | | | | |
| ***GRADE Working Group grades of evidence*** ***High certainty:*** *We are very confident that the true effect lies close to that of the estimate of the effect* ***Moderate certainty:*** *We are moderately confident in the effect estimate: The true effect is likely to be close to the estimate of the effect, but there is a possibility that it is substantially different* ***Low certainty:*** *Our confidence in the effect estimate is limited: The true effect may be substantially different from the estimate of the effect* ***Very low certainty:*** *We have very little confidence in the effect estimate: The true effect is likely to be substantially different from the estimate of effect* | | | | | | | | |

#### *Explanations*

a. *All 12 studies were at risk of performance bias because the participants/parents/clinical team were not masked to intervention. In all except Mazumder's study (weightage 65.4%), the outcome assessors were also not masked to the intervention. However, mortality being a ‘hard’ outcome, we did not downgrade for either performance or outcome assessment bias. Six studies, Acharya, Boo, Cattaneo, Charpak, Eka Prawiti, and Worku contributing to 26.3% weightage in the pooled analysis are at unclear risk of allocation concealment. Four studies Boo, Cattaneo, Suman, and Worku were also at risk of attrition bias due to incomplete outcome data. But they together account for only 22.7% weightage in the pooled analysis. The risk of bias was therefore not downgraded to ‘serious’ risk. One study Mwendwa 2012 was at high risk of bias for random sequence generation and allocation concealment. It contributed to 3.5% weightage.*
b. *All studies were at moderate or severe risk of bias as participants and outcome assessors were not masked to intervention and outcomes. Only in Mazumder, the assessors were masked to the intervention. Though culture-positive sepsis is a ‘hard’ outcome, the largest study Mazumder 2019 that accounted for 91% of weightage defined sepsis based on WHO PSBI signs and not on culture positivity; five studies (4.7% weightage; Ali 2009, Eka Prawiti 2009, Kadam 2005, Kumbhojkar 2016, Suman 2008) did not define sepsis in their studies; another study (Rojas 2008; weightage 9.2%) defined it as both clinical and culture-positive sepsis and only Boo 2007 defined it as culture-positive sepsis. Therefore, the risk of bias was downgraded to ‘serious’ risk. Allocation concealment was unclear in four studies (Charpak 1997, Ali 2009 Boo, 2007 Eka Pratiwi 2009) that together contribute to 4.8% weightage.*
c. *All studies were at high risk of outcome ascertainment bias because the clinical team and outcome assessors were not masked to intervention and outcomes. However, temperature measurement was considered a ‘hard’ outcome. Therefore, we did not downgrade the evidence. Four studies (Acharya, Ali, Eka Prawiti, and Alisjahbana contributing to weightage 32%) were at risk of allocation concealment and two studies (Kumbhojkar and Suman contributing to weightage 23%) were also at risk of incomplete outcome assessment bias. Not downgraded because these studies accounted for <50% weightage.*
d. *Substantial heterogeneity >50%*
e. *All studies were at high risk of outcome ascertainment bias as participants and outcome assessors were not masked to intervention and outcomes. However, weight gain is considered a ‘hard’ outcome. Therefore, we did not downgrade for the risk of bias. Seven studies (Acharya, Ali, Bier, Boo, Cattaneo, Gathwala, and Ramanathan accounting for 64% weightage) were at risk of allocation concealment bias. Therefore, the evidence was downgraded for ‘serious’ risk of bias.*
f. *All studies were at high risk of outcome ascertainment bias because the participants and outcome assessors were not masked to the intervention and the outcome is not a ‘hard’ outcome. Allocation concealment was unclear in 6 studies that accounted for 82% weightage.*
g. *95% confidence intervals overlap no effect (i.e., CI includes RR of 1.0)*
h. *One study Charpak 1997 with moderate risk of bias (unclear allocation concealment; lack of blinding of participants/parents/clinical team and outcome assessors). The follow-up rate at 12-18 months was 80%. The characteristics of infants of KMC and conventional groups who completed follow-up were similar*
i. *Single study*
j. *Only one study with low risk of bias. Developmental outcomes were ascertained in the study clinic by trained psychologists, who were unaware of the group allocation*

**eTable 2: Summary of findings – early-initiated KMC vs. late-initiated KMC infants**

| **Summary of findings Table-2. Early initiated KMC compared to late initiated KMC in preterm or low birthweight infants** | | | | | |
| --- | --- | --- | --- | --- | --- |
| ***Patient or population****: preterm or low birthweight infants*  ***Setting****: Hospital or community/home*  ***Intervention****: Early initiated KMC (within 24 hours after birth)*  ***Comparison****: late initiated KMC (more than 24 hours after birth)* | | | | | |
| ***Outcomes*** | ***№ of participants  (studies) Follow up*** | ***Certainty of the evidence (GRADE)*** | ***Relative effect (95% CI)*** | ***Anticipated absolute effects*** | |
|  |  |  |  | ***Risk with late initiated KMC*** | ***Risk difference with early initiated KMC*** |
| *Mortality by 28 days of age* | *3693 (3 RCTs)* | *⨁⨁⨁⨁ HIGH^a^* | ***RR 0.77*** *(0.66 to 0.91)* | *156 per 1,000* | *36 fewer per 1,000 (53 fewer to 14 fewer)* |
| *Sepsis till 28 days* | *3694 (2 RCTs)* | *⨁⨁◯◯ LOW^b,c^* | ***RR 0.85*** *(0.76 to 0.96)* | *249 per 1,000* | ***37 fewer per 1,000*** *(from 60 fewer to 10 fewer)* |
| *Exclusive breastfeeding - At discharge* | *3464 (3 RCTs)* | *⨁⨁⨁◯ MODERATE^c,d^* | ***RR 1.12*** *(1.07 to 1.16)* | *688 per 1,000* | ***83 more per 1,000*** *(from 48 more to 110 more)* |
| *Exclusive breastfeeding at 28 days of age* | *2841 (3 RCTs)* | *⨁⨁⨁◯ MODERATE^c,d,e^* | ***RR 1.01***  ***(0.98 to 1.04)*** | *855 per 1,000* | ***9 more per 1,000*** *(from 17 fewer to 34 more)* |
| *Hypothermia at discharge or by 28 days* | *3713 (4 RCTs)* | *⨁⨁⨁⨁ HIGH^f^* | ***RR 0.74*** *(0.61 to 0.90)* | *109 per 1,000* | ***28 fewer per 1,000*** *(from 42 fewer to 11 fewer)* |
| *Weight gain at 28 day follow-up (g/d)* | *204 (1 RCT)* | *⨁⨁◯◯ LOW^g,h^* | *-* | *Mean weight gain at 28 day follow-up was* ***12.5*** *g/day* | *MD 2.2 g/day lower (5.26 lower to 0.86 higher)* |
| ******The risk in the intervention group*** *(and its 95% confidence interval) is based on the assumed risk in the comparison group and the* ***relative effect*** *of the intervention (and its 95% CI).*  ***CI:*** *Confidence interval;* ***RR:*** *Risk ratio;* ***MD:*** *Mean difference* | | | | | |

#### *Explanations*

a. *Though parents and the clinical team were not masked to the intervention, mortality was considered a 'hard' outcome, and hence the evidence was not downgraded.*
b. *In both the studies, the participants and clinicians were not masked to the intervention. Both diagnosed sepsis based on WHO's PSBI definition and not by culture positivity. Though the outcome assessment was done by an independent team who was unaware of group allocation in the WHO iKMC study (accounting for 95% of weightage), the risk of performance bias by the clinical team and researchers in a subjective outcome like clinical sepsis or PSBI cannot be ruled out.*
c. *Significant heterogeneity >50%.*
d. *In three studies, participants and clinical team were masked. Assessment of exclusive or any breastfeeding is prone to bias. However, the WHO iKMC contributed to the maximum weightage and the outcome assessment was done by an independent team not involved in the intervention. The risk of performance bias – by the clinical team or researchers – in breastfeeding outcomes was considered low and hence the evidence was not downgraded.*
e. *95% confidence intervals overlap no effect (i.e., CI includes RR of 1.0) but they also exclude important benefit as well as important harm; so not downgraded*
f. *All three studies were at low risk of bias. Although parents and clinical team were not masked to the intervention, measurement of temperature is less prone to outcome assessment bias. Hence not downgraded.*
g. *A single study, that was prematurely terminated at 75% enrollment. We did not downgrade for lack of masking of caregivers or outcome assessors because weight measurement is an objective outcome*
h. *95% CI overlaps no effect (i.e., CI includes RR of 1.0)*
i. *In the WHO iKMC study that accounted for 98% weightage, outcome assessment was done by an independent team whose members were not involved in the intervention delivery. However, hospital discharge was decided by the clinical team who were not masked to intervention and hence the risk of performance bias cannot be ruled out.*
j. *Magnitude of difference in the outcome, despite its statistical significance, is not clinically relevant (~7 hours difference in the hospital stay*

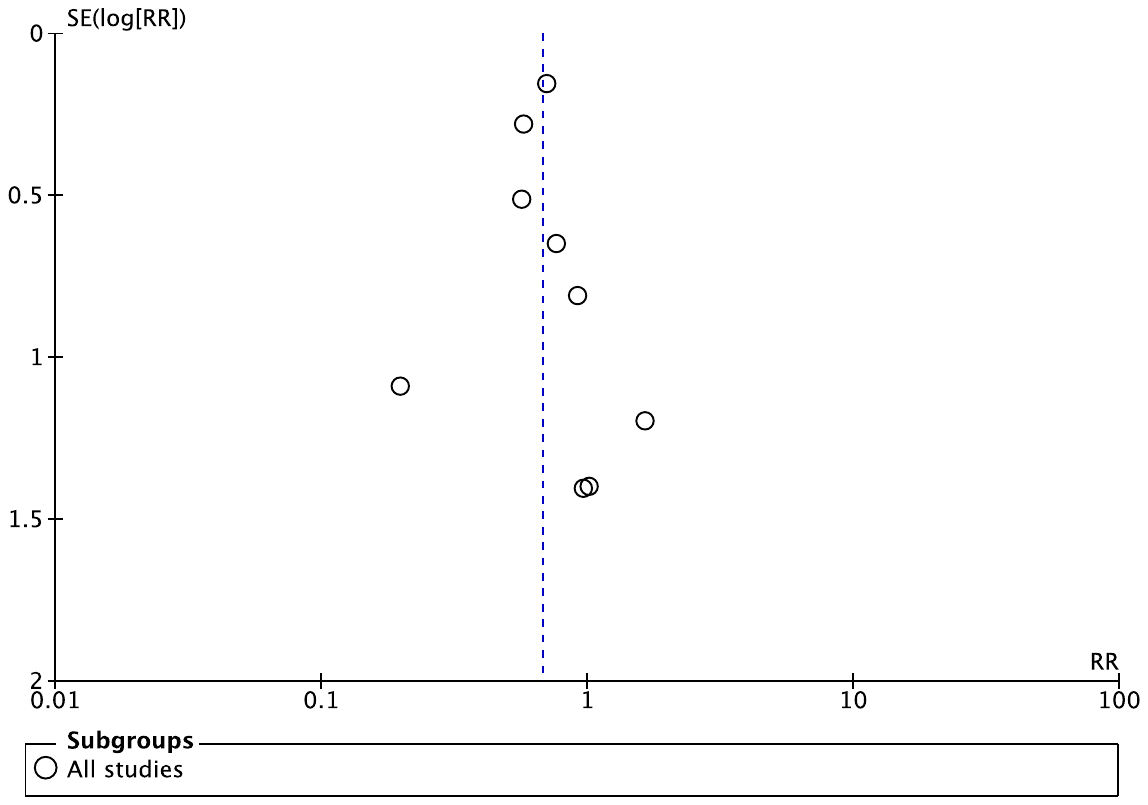

**eFigure 1: KMC vs. conventional care - funnel plot for ‘mortality at discharge/28 days’**

*SE, standard error; RR, relative risk*

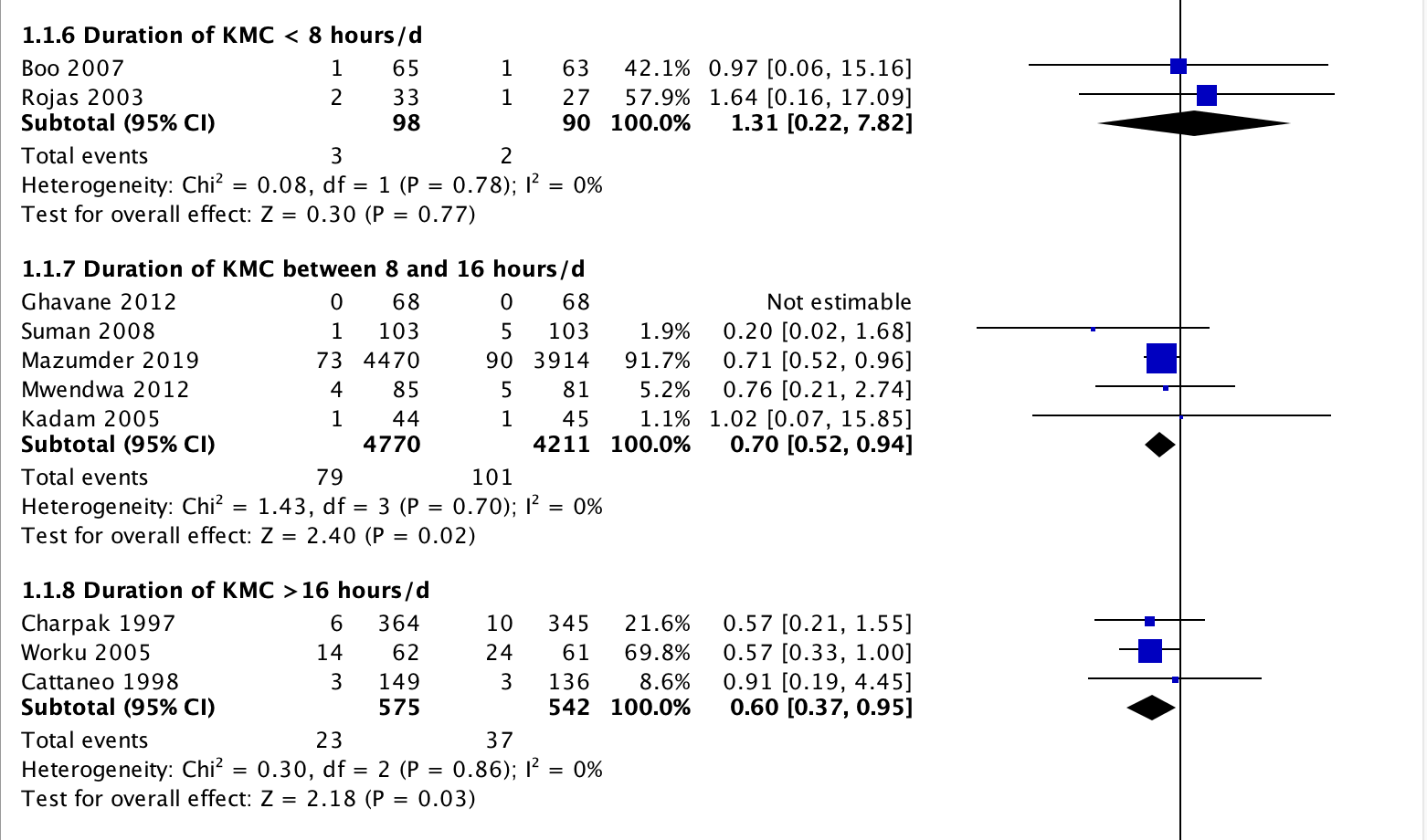

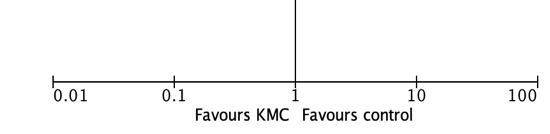

**eFigure 2: KMC vs. conventional care - mortality by discharge/28 days**
(Subgroup analysis by the duration of KMC achieved in the studies)

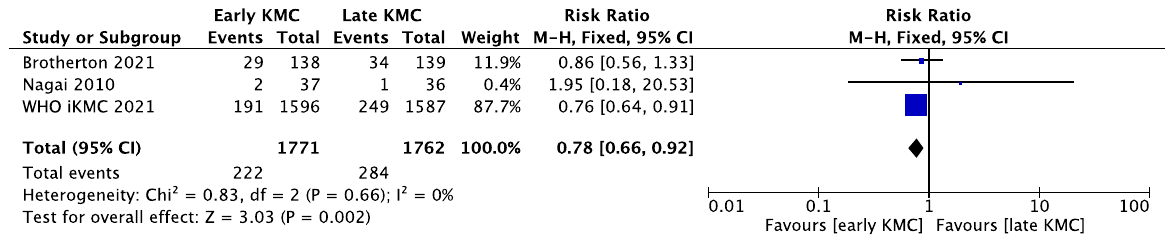

**eFigure 3: Early-initiated vs. late-initiated KMC: mortality by 28 days of life**
